# COVID-19 in Elderly Patients with Acute Kidney Injury

**DOI:** 10.1101/2021.11.16.21266324

**Authors:** Yavuz Ayar, Olgun Deniz, Barıs Doner, Isa Kilic, Canan Demir, Abdulkadir Sahin

**Author notes:** **Corresponding Author:** Yavuz AYAR, University of Health Sciences, Bursa Faculty of Medicine, Department of Nephrology and Internal Medicine, Dogankoy mevki, Nilufer/BURSA. Olgun DENIZ, University of Health Sciences, Bursa Faculty of Medicine, Department of Geriatrics, Dogankoy mevki, Nilufer/BURSA. Baris DONER, University of Health Sciences, Bursa Faculty of Medicine, Department of Nephrology, Dogankoy mevki, Nilufer/BURSA. Isa KILIC, University of Health Sciences, Bursa Faculty of Medicine, Department of Intensive Care, Anesthesia and Reanimation, Dogankoy mevki, Nilufer/BURSA. Canan DEMIR, University of Health Sciences, Bursa Faculty of Medicine, Department of Clinical Microbiology and Infection Diseases, Dogankoy mevki, Nilufer/BURSA. Abdulkadir SAHIN, University of Health Sciences, Bursa Faculty of Medicine, Department of Internal Medicine, Dogankoy mevki, Nilufer/BURSA.

## Abstract

**Objective:** Coronavirus disease 2019 (Covid 19) started in China in December 2019 and spread all over the world, is more progressive in patients who are elderly and with chronic diseases. Especially kidney involvement affects the survival of patients. In this study, we analyzed Covid 19 patients who developed acute kidney injury treated in our unit, retrospectively.

**Matherials:** The clinical and laboratory data of 610 patients who hospitalized due to Covid 19 pandemic between 01.06.2020 and 30.06.2021 in the intensive care and other clinics of our hospital evaluated from the records, retrospectively. One hundred-fourty patients diagnosed with AKI according to the criteria of KDIGO (Kidney Disease Global Outcomes). The patients divided into two groups as KDIGO stage 1 and 2, 3.

**Results:** The median age in both groups was 70 (35-92) and 73 (35-90) years. Approximately seventy percent of them were over 65 years old. Almost all of the patients had hypertension. Most of the patients were using angiotensin converting enzyme inhibitors (ACE inh) or angiotensin receptor blockers (ARB) (84%). AKI was present at the time of admission (61.9%) in the KDIGO 1 group and at the time of hospitalization (64.3%) in the KDIGO 2, 3 group. The mortality rate was higher in stage 2-3 AKI patients (35.7%). Ferritin and fibrinogen levels were high in the KDIGO 2, 3 group, while lymphocyte levels were low.

**Conclusion:** AKI can be seen at the time of admission and during treatment in patients who are hospitalized and treated due to Covid 19. Covid 19 is more mortal in patients with advanced AKI.

## INTRODUCTION

Coronavirus disease 2019 (Covid 19) started to affect the world with the end of 2019. As a global pandemic, it resulted in more than 175 million cases and approximately 3.8 million deaths as of June 13 (1). Acute kidney injury (AKI) in Covid 19 patients varies from country to country, from region to region, but is observed between 10-15%, and this rate reaches up to 50% in patients hospitalized in the intensive care unit. AKI has a serious course especially in patients hospitalized in the intensive care unit, leading to prolonged hospitalizations and increased mortality (2-4).

The aim of our study was to evaluate the clinical and laboratory data, risk factors affecting prognosis and mortality of patients hospitalized for AKI in a tertiary healthcare institution.

## MATHERIALS and METHODS

### Patients

The data of 610 patients over the age of 18 who admitted to our hospital and hospitalized in intensive care unit and clinics between 01.06.2020 and 30.06.2021 due to Covid 19 evaluated retrospectively. According to Kidney Disease Global Outcomes (KDIGO) AKI guidelines (Table 1), patients diagnosed with AKI and divided into two groups as KDIGO 1 and KDIGO 2, 3. The clinical and laboratory data of the patients analyzed.

**Table 1.** Staging of acute kidney injury according to KDIGO guideline (5).

|  |  |  |
| --- | --- | --- |
| Stage 1 | Serum creatinine | Urine output |
| 1 | 1.5–1.9 times baseline OR $\geq 0.3$ mg/dl<br>($\geq 26.5$ mmol/l) increase | <0.5 ml/kg/h for 6– 12 hours |
| 2 | 2.0–2.9 times baseline | <0.5 ml/kg/h for $\geq 12$ hours |
| 3 | 3.0 times baseline | <0.3 ml/kg/h for $\geq 24$ hours |
|  | OR | OR |
| | Increase in serum creatinine to 4.0 mg/dl ($\geq 353.6$ mmol/l) | Anuria for $\geq 12$ hours |
|  | OR |  |
|  | Initiation of renal replacement therapy |  |
|  | OR, In patients <18 years, decrease in eGFR to <35 ml/min per 1.73 m<sup>2</sup> |  |

### Biostatistical analysis

Statistical analysis was carried out using the Statistical Package for the social sciences (SPSS) version 21.0 (IBM, Armonk, NY, USA) software program. Continuous variables were assessed by Kolmogorov-Smirnov test and histograms to find out if they had normal or skewed distribution. Continuous skew-distributed variables were compared with the Mann– Whitney *U-*test between the two groups, and variables with normal distribution were compared with independent variables *t*-test between the two groups. Categorical variables were compared by Chi-square or Fisher Exact tests, where appropriate. Categorical variables were presented as number and frequency. P-value < 0.05 was considered statistically significant.

## RESULTS

Approximately 23% (140 patients) of 610 patients admitted to the hospital diagnosed with AKI according to the KDIGO AKI guidelines. Most of the patients defined as stage 1 AKI (80%). The number of patients over 65 years of age in both groups was 67.9%. As an additional disease, 97% of AKI patients had hypertension. Eighty four percent of the patients were using angiotensin converting enzyme inhibitors (ACEinh) or angiotensin receptor blockers (ARB) as antihypertensive therapy, 75% of the patients were using antiaggregant or anticoagulant therapy. The number of patients with diabetes (39.3%) and using insulin (28.6%) were higher in the KDIGO 1 group. The duration of AKI was 5 (2-25) days in the KDIGO 1 group and 7 (2-25) days in the KDIGO 2, 3 group and did not differ. AKI development during hospitalization was higher in the KDIGO 2,3 group (61.9%), and AKI at admission was more common in the KDIGO 1 group (64.3%) (p=0.003 and 0.013, respectively). Mortality was 35.7% higher in the KDIGO 2.3 group (p=0.001). Ferritin and fibrinogen levels, as markers of infection and inflammation, increased more in the KDIGO 2, 3 group (p=0.042 and 0.017, respectively). Lymphocyte count and oxygen saturation were lower in the same group (p<0.001 and 0.002, respectively). Clinical and laboratory findings are summarized in Table 2.

**Table 2.** Characteristics and laboratuary findings in both groups.

|  | KDIGO stage 1<br>(n=112) | KDIGO stage 2 and 3<br>(n=28) | p |
| --- | --- | --- | --- |
| Age, median (min-max) | 70 (35-92) | 73 (35-90) | 0.630 |
| Age ≥ 65, n (%) | 76 (67.9) | 76 (67.9) | 1 |
| Diabetes Mellitus, n (%) | 44 (39.3) | 8 (28.6) | 0.294 |
| Hypertension, n (%) | 109 (97.3) | 27 (96.4) | 0.800 |
| Chronic Kidney Disease, n (%) | 26 (23.2) | 6 (21.4) | 0.840 |
| Obesity, n (%) | 2 (1.8) | 1 (3.6) | 0.491 |
| Chronic Obstructive Pulmonary Disease, n (%) | 16 (14.3) | 3 (10.7) | 0.765 |
| Coronary Artery Disease, n (%) | 46 (41.1) | 11 (39.3) | 0.863 |
| Heart Failure, n (%) | 33 (29.5) | 4 (14.3) | 0.103 |
| Cerebrovasculer Disease, n (%) | 6 (5.4) | 3 (10.7) | 0.383 |
| Malignity, n (%) | 13 (11.6) | 7 (25) | 0.126 |
| Chronic liver disease, n (%) | 2 (1.8) | 0 (0) | 0.476 |
| Medications |  |  |  |
| -ACE inh, n (%) | 54 (48.2) | 13 (46.4) | 0.866 |
| -ARB, n (%) | 41 (36.6) | 10 (35.7) | 0.930 |
| -CCB, n (%) | 42 (37.5) | 10 (35.7) | 0.861 |
| -BB, n (%) | 72 (64.3) | 13 (46.4) | 0.084 |
| -Insulin, n (%) | 32 (28.6) | 3 (10.7) | 0.051 |
| -OAD, n (%) | 13 (11.6) | 5 (17.9) | 0.359 |
| -Antiagregan, n (%) | 78 (69.6) | 16 (57.1) | 0.208 |
| -Anticoagulan, n (%) | 9 (8) | 2 (7.1) | 0.875 |
| Duration of acute kidney injury, n (%) | 5 (2-25)<br>(n =39) | 7 (2-25)<br>(n =16) | 0.386 |
| Acute renal failure |  |  |  |
| -during hospitalization, n (%) | 38 (33.9) | 18 (64.3) | **0.003** |
| -at admission, n (%) | 69 (61.9) | 10 (35.7) | **0.013** |
| AKI on CKD | 26 (23.2) | 6 (21.4) | 0.626 |
| AKI progression, n (%) | 10 (8.9) | 11 (39.3) | **<0.001** |
| Mortality, n (%) | 9 (8) | 10 (35.7) | **0.001** |
| Duration of intensive care unit, median (min-max) | 8 (2-45) | 6 (1-21) | 0.546 |
| Ferritin (µg/l) | 304.10 (23.40 – 2000) | 517.50 (74.10 – 2000) | **0.042** |
| Lenfosit (10<sup>3</sup>/µl) | 1.32 (0.24 – 4.92) | 0.89 (0.06 – 2.13) | **< 0.001** |
| D dimer (µg/ml) | 9.50 (1.5-92.2) | 10.2 (2-91.6) | 0.306 |
| Fibrinogen (mg/dl) | 424 (148-900) | 511.50 (189-900) | **0.017** |
| Serum creatinine (mg/dl) | 1.47 (0.56-5.35) | 1.43 (0.56-8.38) | 0.942 |
| Serum Na (mmol/l) | 137 ± 4.9 | 136 ± 4.9 | 0.206 |
| Serum albumin (g/dl) | 3.45 ± 0.47 | 3.32 ± 0.44 | 0.166 |
| CRP level, ≥5, n (%) | 98 (87.5) | 27 (96.4) | 0.304 |
| Oxygen saturation > 90, n (%) | 97 (86.6) | 17 (60.7) | **0.002** |

## DISCUSSION

AKI was the common outcome renal disease reported by studies in COVID-19, particularly among patients in the intensive care unit (ICU). The reported rates of AKI are extremely variable; however, available evidence suggests that it likely affects 0.5 to 40% of hospitalized patients and 50 to 80% of patients in the ICU (6-8).

Histopathological data relating to COVID-19 AKI are limited. The most common cause of acute kidney injury is seen as acute tubular injury in postmortem studies. Virus particles also damage tubular epithelium and podocyte. Importantly, complement activation and thrombotic microangiopathy are important mechanisms of kidney injury. COVID-19 is also associated with activation of an inflammatory response that has been termed a ‘cytokine storm’, which might contribute to the pathogenesis of COVID-19-associated multi-organ dysfunction. In studies reported that the virus causes collapsing glomerulopathy, which is a subtype of FSGS (in those with APOL1 gene variant), and glomerular diseases such as immunocomplex, ANCA-associated glomerulonephritis and IgA nephropathy. Activation of the renin-angiotensin aldosterone system, hyperinflammatory response, hypercoagulability, and nonspecific factors such as hypotension and hypoxemia facilitate the development of AKI. Management of the disease in hospital and intensive care unit (dehydration, secondary infection, sepsis, etc.) also facilitates the development of acute kidney injury (9-13). In our study, sepsis (increased ferritin levels and lymphopenia) and hypoxemia were more prominent in the KDIGO 2,3 group.

Older age, chronic kidney disease (CKD), and the presence of other comorbidities (for example, diabetes mellitus, hypertension, obesity, heart failure and chronic obstructive pulmonary disease) are associated with worse outcomes and also represent risk factors for the development of AKI in patients with COVID-19. Renal replacement therapy used in 5% to 11% in hospitalized patients and 16% to 50% in ICU (14, 15). In our study, most of our patients had hypertension as a comorbid disease. However, 23.2% of KDIGO 1 group and 21.4% of KDIGO 2, 3 group had chronic kidney disease (acute kidney injury over chronic kidney disease). The need for renal replacement therapy developed in 39.3% of the patients in the KDIGO 2,3 group and 8.9% in the KDIGO 1 group. AKI progression was more evident in the KDIGO 2.3 group.

Mortality rate can be high in Covid 19 AKI patients. Studies shew that the mortality rate in Covid 19 KDIGO 2,3 AKI patients detected 11.2-91.7% (2, 3). We found AKI in 23% of our patients. The mortality rate was higher in these patients (13.6%), especially in the KDIGO 2, 3 group. Early diagnosis of the disease, hospitalization, comorbidities and management of critically ill patients affect mortality. In our country, effective treatment and hospitalization, efficient use of resources led to lower mortality rates.

In conclusion, COVID 19 affects many organ systems. The progression of the disease is especially serious in organ involvement such as renal involvement. AKI is the most common kidney disease in the course of the disease. Mortality is observed more in advanced AKI. Early diagnosis, effective treatment, hospitalization and follow-up in severe patients are important in the treatment of the disease.

## Data Availability

All data produced in the present study are available upon reasonable request to the authors

## DECLERATIONS

### Source of funding

None

